# Association between Abnormal Fetal Head Growth and Autism Spectrum Disorder

**DOI:** 10.1101/2020.08.09.20170811

**Authors:** Ohad Regev, Gal Cohen, Amnon Hadar, Jenny Schuster, Hagit Flusser, Analya Michaelovski, Gal Meiri, Ilan Dinstein, Reli Hershkovitch, Idan Menashe

## Abstract

Despite evidence for prenatal onset of abnormal head growth in children with autism spectrum disorder (ASD), fetal ultrasound studies in ASD are limited and controversial. We conducted a longitudinal matched case-sibling-control study on fetal ultrasound biometric measures from 174 ASD children, their own typically developed siblings (TDS; n=178) and other population-based typically developed children (TDP; n=176). During second trimester, ASD and TDS fetuses had significantly smaller biparietal diameter (BPD) than TDP fetuses (aOR_zBPD_=0.685, 95%CI=0.527-0.890 and aOR_zBPD_=0.587, 95%CI=0.459-0.751, respectively). Interestingly, sex had a significant effect on head growth with males having larger heads than females within and across groups. Also, males and females with ASD showed different head shapes which were inversely correlated with ASD severity across different gestation periods. Our findings suggest that abnormal fetal head growth is a familial trait of ASD, which is modulated by sex and is associated with the severity of the disorder.

## Introduction

Autism spectrum disorder (ASD) is a life-long neurodevelopmental disorder with a complex etiology having both genetic and environmental determinants.^1,2^ The wide range of pre- and perinatal risk factors,^3,4^ together with the abnormalities in early brain development associated with ASD,^5-10^ suggest that, at least in some cases, the predisposition to ASD begins during gestation.

Abnormal head growth has been proposed as an early biomarker for ASD. Specifically, various MRI, postmortem, and head measurement studies have reported that infants who later developed ASD were born with a smaller or average head circumference (HC), followed by an excessive increase during the first years of life in head size, in extra-axial cerebrospinal fluid (CSF) volume, and in brain regions associated with the language, social, emotional and communication functions affected by ASD.^7,8,19,20,11-18^ These studies also reported correlations between clinical severity to postnatal head size, brain morphometry and extra-axial CSF volume.^15,19,21,22^

Importantly, some of these head growth abnormalities have been associated with sex, which is significantly associated with ASD, with the disorder being ~4 times more prevalent among males, but symptoms tending to be more frequent and severe among women.^23-27^Several postnatal studies have found sex- specific neuroanatomical profiles in ASD children compared to sex-matched controls. Among these investigations, a large population-based study revealed sex-specific differences in HC at birth and throughout early childhood in children with ASD.^18^ Similarly, MRI studies found sex-specific differences in total brain volume (in gray and white matter volume of the frontal and temporal lobes, cingulate cortex, amygdala, and cerebellum), alongside different and abnormal growth trajectories throughout childhood.^14,16,22,28-30^ These studies also showed sex-specific correlations between specific brain regions and the clinical severity of ASD.^22^

Previous postnatal studies suggested that the accelerated head growth among ASD children during early childhood may be a reaction or an adaptation to abnormal prenatal head growth. This assertion is supported by an emerging ASD prenatal literature covering developmental abnormalities in certain fetal ultrasound (US) parameters,^31,32^ and specifically in biometric measures.^33-37^ Nevertheless, these studies showed conflicting results. Two studies found indications for either wider heads with normal HC in ASD children compared to typically developed population (TDP) controls^33^ or larger HC with normal shape compared to population norm.^34^ Two other studies found an association between smaller HC to either ASD phenotype^36^ or autistic traits^37^ while the fifth study didn’t find any association between HC and ASD.^35^ In addition, the reported studies were conducted on relatively small cohorts and did not account for potential confounders, e.g., none took into consideration familial factors known to play a significant role in fetal growth^38^ and/or to constitute a risk for ASD.^39^ Similarly, several studies did not take into account the inherent male sex bias in ASD,^33,34^ even though boys are known to have larger biometric measures.^40^ Finally, these studies report findings from different gestational periods, which further complicates comparing their findings. Thus, conclusions regarding fetal head growth in children with ASD remain minimal, inconsistent and controversial. The main goal of this study, was therefore, to assess fetal head growth in fetuses later diagnosed with ASD in comparison to their typically developed siblings and to a population control across different gestation periods. In addition, we asked whether differences in fetal head growth are modulated by the sex of the fetus and whether they are associated with the clinical severity of children with ASD.

## Methods

### Study Sample

The participants in this case/control study were singletons whose families were members of Clalit Health Services (CHS), Israel’s largest health maintenance organization, serving about 75% of the approximately 700,000 residents of southern Israel, composed of Jews and Bedouins, two ethnic groups that differ in their genetic background and environmental exposures. Members of CHS in this region receive most of their hospital-related services (including ASD diagnosis) at the Soroka University Medical Center (SUMC), the region’s only tertiary hospital. The diagnosis of ASD at SUMC is a multidisciplinary process, which entails a comprehensive intake interview (clinical and socio-demographic factors), a behavioral evaluation with ADOS-2,^41^ and a full neurocognitive assessment (e.g., Bayley-III^42^ or WPPSI-III^43^). Final diagnoses of ASD is made by a pediatric psychiatrist or neurologist, according to DSM-5 criteria.^26^ All sociodemographic and clinical data on these children are stored in the database of the National Autism Research Center of Israel (NARC).^44^

A flowchart showing the case/control assignment for this study is presented in **Figure 1**. We randomly selected from the NARC database 386 of the 719 children diagnosed with ASD (53.7%). Among these 386 cases, we identified those for whom there were prenatal US scans in the CHS database. The 212 children (54.9%) for whom US scans were missing were excluded from the study, leaving us with a sample of 174 cases, born between 2008-2017. There were no significant sociodemographic or clinical differences between this sample and other children with ASD in the NARC database, except that the proportion of Jews (vs. Bedouin Israelis) was slightly lower for children included in the study, as was the parental age (**Table 1**).

**Figure 1.**
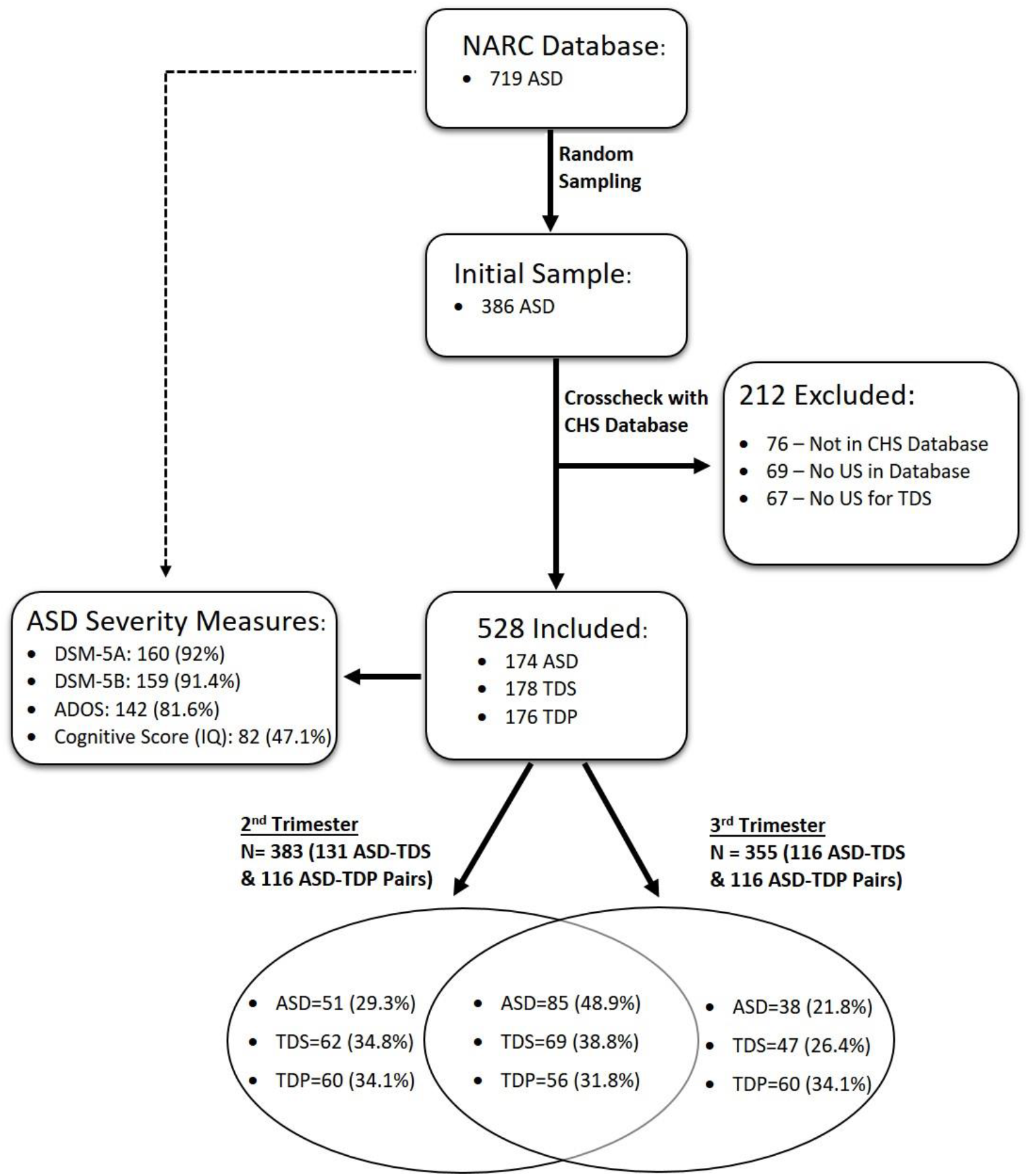
Flowchart of Children included in this Study.

**Table 1.** Clinical and Sociodemographic Characteristics of ASD Children.

| Variable | Included<br>(n = 174) | Excluded<br>(n = 545) | P-Value |
| --- | --- | --- | --- |
| Jewish, no. (%) <sup>a</sup> | 124(72.1) | 427 (79.8) | **0.034** |
| Male, no. (%) <sup>a</sup> | 142(82.1) | 418(76.7) | 0.136 |
| Moderate salary or lower, No. (%) <sup>a</sup> | 30(61) | 104(63.8) | 0.743 |
| Special education setting, No. (%) <sup>a</sup> | 47(42) | 136(43) | 0.844 |
| Mother's age, mean (SD), y <sup>b</sup> | 29.3(5.4) | 31.5(6.2) | **<0.001** |
| Father's age, mean (SD), y <sup>b</sup> | 32.6(7.4) | 34.7(7.6) | **0.005** |
| Diagnosis age, mean (SD), y <sup>b</sup> | 3.1(1.2) | 3.2(1.5) | 0.639 |
| Cognitive score (IQ), mean (SD) <sup>b</sup> | 78.4(13.3) | 75.5(17.2) | 0.112 |
| ADOS Comparison Score, median (IQR) <sup>c</sup> | 8(6-9) | 7(6-9) | 0.500 |

<sup>a</sup>Chi-square; <sup>b</sup>Two-sided t-test; <sup>c</sup>Mann-Whitney U test

Of the 174 cases included in the study, 85 had US scans from both the second and third trimesters, while 51 and 38 had scans from only the second or the third trimester, respectively. For each trimester, we matched two types of control group for cases: (1) *typically developing siblings (TDS) -* for each ASD case, we matched her/his own typically developed sibling who was closest in age; and (2) *typically developed population (TDP) -* for each case, we identified a typically-developed child matched by year of birth, sex and ethnicity. In total, the study sample included 528 children: 174 with ASD, 178 TDS, and 176 TDP (**Figure 1**).

### Fetal Ultrasound Data

All the US scans had been performed by experienced sonographers or physicians that recorded fetal biometric measures using standardized US measurements of simple anatomic landmarks to the nearest millimeter. The following biometric measures were extracted from the CHS database for each trimester: HC and head width (BPD, biparietal diameter), i.e., two biometric measures that strongly correlate with brain size;^45,46^ abdominal circumference (AC); and femur length (FL). HC was measured at the level of the BPD, taken at the widest point of the skull at the level of the thalamic peduncles and the cavum septum pellucidum; BPD was measured from outer skull wall to the inner wall directly opposite; AC was measured at the level of the junction of the vertebra, portal sinus, and fetal stomach; and FL was measured from the greater trochanter to the distal metaphysic.^33,47,48^ It is known that for these US measurements there is very high inter-rater reliability between experienced sonographers, particularly when transformation to standardized Z-scores are used.^33,36,49,50^ Gestational age (GA) of each child was calculated from the last menstrual period (LMP) and confirmed by the crown-rump length (CRL) from the first trimester. If the date of LMP was unknown, GA was calculated based on CRL.

### Statistical Analysis

We converted all biometric fetal measures to gestation-matched standardized Z-scores using the Hadlock approach,^48,51-53^ the most widely used standardization approach in this field. In addition, we created five new variables by subtracting body parameter (AC and FL) Z-scores from head parameter (HC and BPD) Z-scores and by subtracting the HC Z-scores from the BPD Z-scores to assess fetal head growth relative to the other biometric measures.^33,35^ If there was more than one US measure for a fetus in a particular trimester, we used the mean of its Z-scores as a representative measure. Notably, biometric measures with Z-scores > ±6 or that deviated by >3 SD from the average for that trimester were considered outliers and removed from further analyses (29 outliers: 12 ASD, 9 TDP, 8 TDS).

Differences in sociodemographic characteristics between cases and the two control groups were assessed by using the appropriate univariate statistics. Differences in biometric measures between cases and each of the two matched control groups (TDS and TDP) were assessed using paired t-tests and multivariable conditional regression adjusting for sex and maternal age. Differences in biometric measures between the two matched control groups were assessed via unpaired t-tests and multivariable unconditional logistic regression adjusting for sex, maternal age and ethnicity. For fetuses with longitudinal data, a linear mixed-effect model was used to assess the association between fetal growth rate (measured as the difference in biometric measures throughout pregnancy) and ASD, taking into account repeated biometric measures, ethnicity and gestational age.^36^ These analyses were conducted separately for males and females to examine the effect of sex on fetal growth rate. Lastly, correlation coefficients were calculated in cases between biometric measures and ASD severity according to ADOS-2, DSM-5 and cognitive measures. P-values of analyses with multiple testing were adjusted using the Bonferroni correction. All analyses were conducted using SPSS Statistics V. 25 and R software. A two-sided test significance level of 0.05 was used throughout the entire study.

## Results

### Sociodemographic and Clinical Characteristics

Clinical and sociodemographic characteristics of the study sample are shown in **Table 2**. As expected, the sex ratio for the TDS group was significantly different from that for the ASD group, close to 1:1 vs 4:1 male/female, respectively (p <0.001). In addition, for the TDP group, US scans were performed earlier in the second trimester compared to cases (21.4±2.8 vs. 22.3±1.8 weeks, respectively; p=0.006). No other statistically significant differences were seen between cases and controls.

**Table 2.** Clinical and Sociodemographic Characteristics for Children included in this Study.

| Variable |  | ASD (n = 174) | TDS (n = 178) | TDP (n = 176) |
| --- | --- | --- | --- | --- |
| Ethnicity (Jewish) | No. (%) | 127(73) | 131(73.6) | 139(79) |
|  | P-value<sup>a</sup> |  | 1 | 0.380 |
| Sex (male) | No. (%) | 140(80.5) | 98(55.1) | 146(83) |
|  | P-value<sup>a</sup> |  | **<0.001** | 1 |
| Pregnancy number | Median (IQR) | 2(1-3) | 2(2-4) | 2(1-3) |
|  | P-value<sup>d</sup> |  | 0.208 | 0.172 |
| Previous abortions | Median (IQR) | 0(0-1) | 0(0-1) | 0(0-1) |
|  | P-value<sup>d</sup> |  | 1 | 0.312 |
| C section | No. (%) | 31(18.5) | 23(13.4) | 26(17.9) |
|  | P-value<sup>a</sup> |  | 0.400 | 1 |
| Age at birth (weeks) | Mean (SD) | 38.8(2.5) | 39.0(2) | 39.3(1.4) |
|  | P-value<sup>c</sup> |  | 1 | 0.164 |
| Birth weight (g) | Mean (SD) | 3174 (579) | 3186(552) | 3324(523) |
|  | P-value<sup>c</sup> |  | 1 | 0.052 |
| 1-min low APGAR score (<7) | No. (%) | 4(5) | 5(5) | 6(6) |
|  | P-value<sup>b</sup> |  | 1 | 1 |
| Mother's age (years) | Mean (SD) | 28.6(5.8) | 28.6(5.2) | 29.2(4.8) |
|  | P-value<sup>c</sup> |  | 1 | 0.510 |
| Examined by physician | No. (%) | 136(85) | 140(78.7) | 90(79.6) |
|  | P-value<sup>a</sup> |  | 0.264 | 0.496 |
| Breech Presentation at US | No. (%) | 51(30.9) | 39(22.4) | 40(24.1) |
|  | P-value<sup>a</sup> |  | 0.334 | 0.308 |
| Gestation age assessed by last menstrual period | No. (%) | 115(74.7) | 129(75.9) | 85(79.4) |
|  | P-value<sup>a</sup> |  | 1 | 0.742 |
| Gestational age at 2<sup>nd</sup> trimester US (weeks) | Mean (SD) | 22.3(1.8) | 22.5(1.5) | 21.4(2.8) |
|  | P-value<sup>c</sup> |  | 0.632 | **0.006** |
| Gestational age at 3<sup>rd</sup> trimester US (weeks) | Mean (SD) | 32.5(2.6) | 32.6(2.4) | 33.2(2.9) |
|  | P-value<sup>c</sup> |  | 1 | 0.132 |

<sup>a</sup> Chi-square; <sup>b</sup> Fisher's Exact Test; <sup>c</sup> Two-sided t-test; <sup>d</sup> Mann-Whitney U test . All p-values are Bonferroni corrected for multiple comparison ( $n=2$ ).

### Biometric Measures

Differences in fetal US biometric measures between cases and controls are shown in **Table 3** and **Figures 2&3**. In the second trimester, both ASD and TDS fetuses had significantly narrower heads (smaller BPD) than TDP fetuses (aOR_zBPD_=0.685, 95%CI=0.527-0.890 and aOR_zBPD_=0.587, 95%CI =0.459-0.751, respectively), giving the head a dolichocephalic shape. Interestingly, the BPD of the TDS controls was the smallest of all groups, leading to a smaller HC, which was also significantly smaller than the HC for the TDP group (a0RzHc=0.598, 95%CI=0.426-0.838). The heads of ASD and TDS fetuses were also proportionally smaller vs other body parts compared to TDP fetuses (**Table 3**), indicating that these differences are specific to the head and are not due to differences in fetal growth or fetal age. By the third trimester, these differences in biometric measures between groups had attenuated and had become statistically indistinguishable, although the BPD of the ASD fetuses had become the smallest of all groups (**Table 3**).

**Figure 2:**
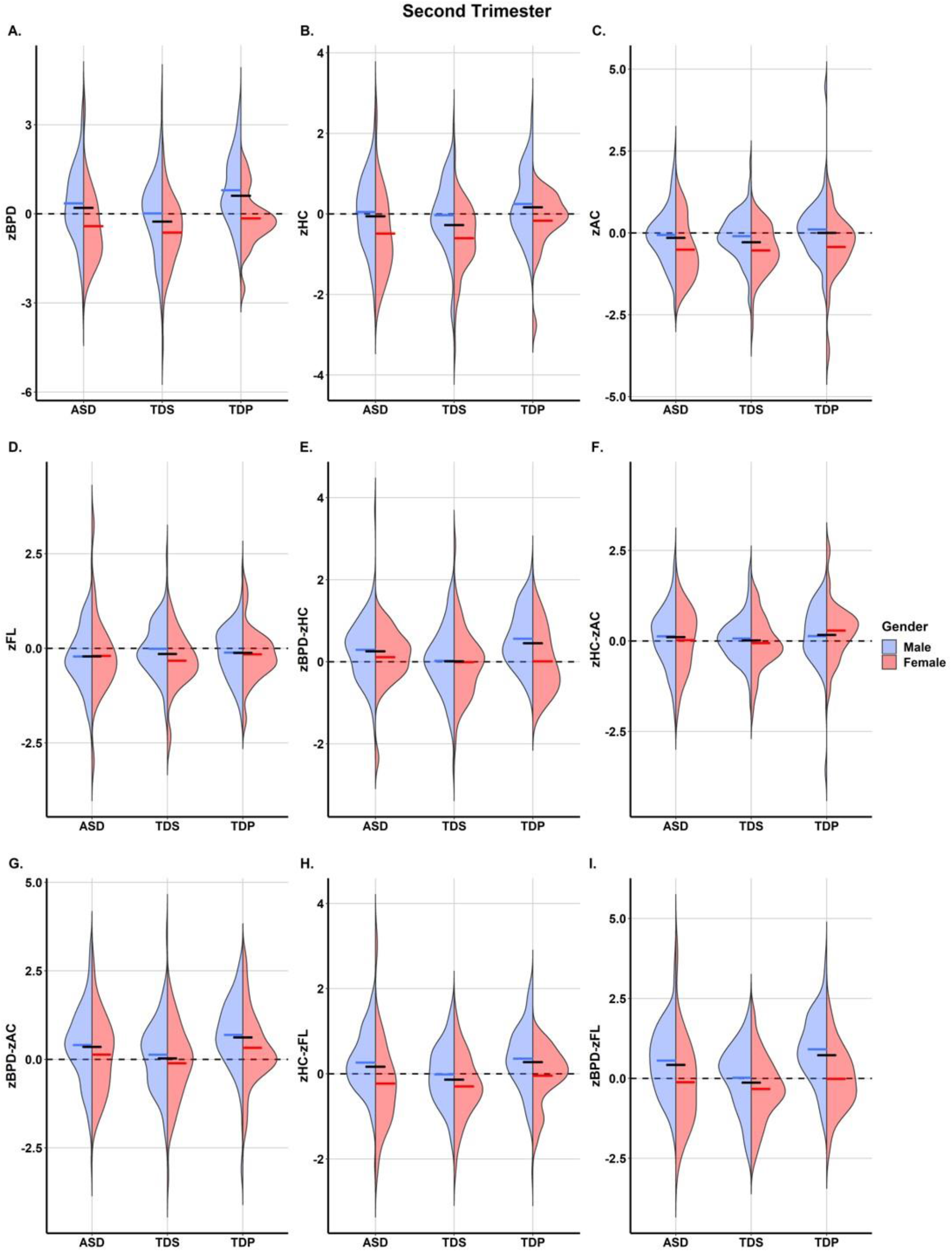
Half-Violin Plots of the Biometric Measures during the Second Trimester. In each panel the density (violin shape) and mean (horizontal lines) of a different biometric measure is shown for males (blue) and females (red) in the ASD, TDS and TDP groups. Black horizontal lines represent the mean measures for each group.

**Figure 3:**
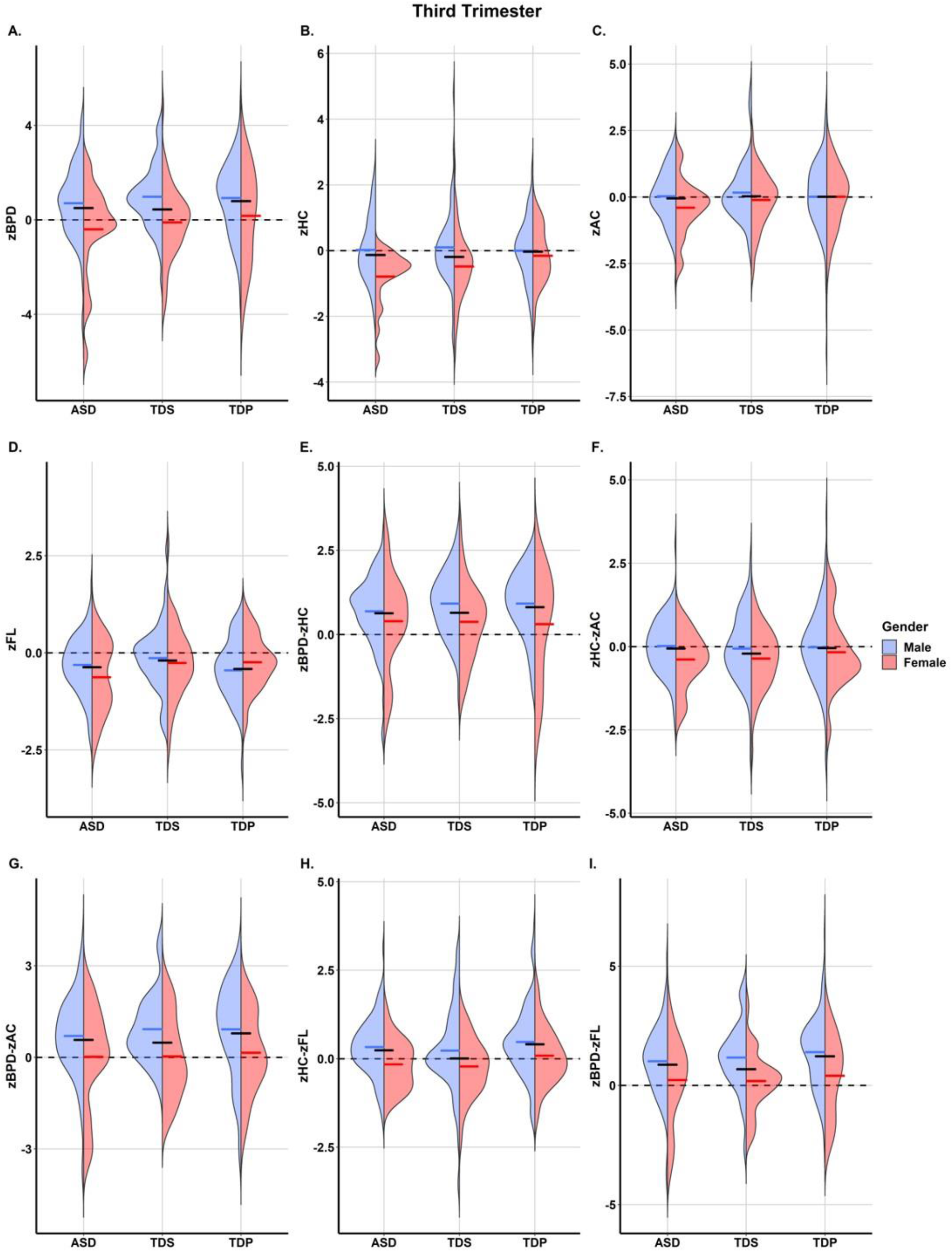
Half-Violin Plots of the Biometric Measures during the Third Trimester. In each panel the density (violin shape) and mean (horizontal lines) of a different biometric measure is shown for males (blue) and females (red) in the ASD, TDS and TDP groups. Black horizontal lines represent the mean measures for each group.

**Table 3.** Group Differences in Fetal Biometric Measures – All Children.

| Variable | Group comparison | Mean difference ± SD | P-value<sup>a</sup> | Adjusted odds ratio (aOR)<sup>b</sup> | 95% CI | P-value<sup>a</sup> |
| --- | --- | --- | --- | --- | --- | --- |
| **Second Trimester: 131 ASD-TDS pairs, 116 ASD-TDP pairs** |  |  |  |  |  |  |
| zBPD | ASD(Ref) - TDS | 0.420±1.63 | **0.012** | 1.261 | 0.998-1.594 | 0.156 |
| | ASD(Ref) - TDP | -0.402±1.53 | **0.018** | 0.685 | 0.527-0.890 | **0.015** |
| | TDS(Ref) - TDP | -0.870±1.74 | **<0.001** | 0.587 | 0.459-0.751 | **<0.001** |
| zHC | ASD(Ref) - TDS | 0.181±1.24 | 0.300 | 1.087 | 0.795-1.487 | 1 |
| | ASD(Ref) - TDP | -0.216±1.11 | 0.12 | 0.693 | 0.486-0.987 | 0.126 |
| | TDS(Ref) - TDP | 0.441±1.26 | **<0.001** | 0.598 | 0.426-0.838 | **0.009** |
| zAC | ASD(Ref) - TDS | 0.094±1.09 | 0.984 | 0.991 | 0.701-1.400 | 1 |
| | ASD(Ref) - TDP | -0.180±1.27 | 0.402 | 0.809 | 0.595-1.100 | 0.531 |
| | TDS(Ref) - TDP | -0.285±1.25 | **0.036** | 0.792 | 0.571-1.097 | 0.480 |
| zFL | ASD(Ref) - TDS | -0.071±1.07 | 1 | 0.838 | 0.596-1.177 | 0.921 |
| | ASD(Ref) - TDP | -0.099±1.04 | 0.933 | 0.846 | 0.591-1.212 | 1 |
| | TDS(Ref) - TDP | -0.026±1.02 | 1 | 1.061 | 0.733-1.536 | 1 |
| zBPD-zHC | ASD(Ref) - TDS | 0.257±1.13 | **0.033** | 1.577 | 1.081-2.302 | 0.054 |
| | ASD(Ref) - TDP | -0.181±0.96 | 0.141 | 0.621 | 0.407-0.948 | 0.081 |
| | TDS(Ref) - TDP | -0.442±1.12 | **<0.001** | 0.521 | 0.364-0.746 | **<0.001** |
| zHC-zAC | ASD(Ref) - TDS | 0.101±1.06 | 0.837 | 1.159 | 0.811-1.657 | 1 |
| | ASD(Ref) - TDP | -0.025±1.16 | 1 | 0.939 | 0.679-1.298 | 1 |
| | TDS(Ref) - TDP | -0.153±1.10 | 0.369 | 0.715 | 0.498-1.027 | 0.210 |
| zBPD-zAC | ASD(Ref) - TDS | 0.324±1.41 | **0.027** | 1.369 | 1.041-1.800 | 0.072 |
| | ASD(Ref) - TDP | -0.238±1.49 | 0.276 | 0.767 | 0.588-1.000 | 0.150 |
| | TDS(Ref) - TDP | -0.592±1.46 | **<0.001** | 0.572 | 0.431-0.758 | **<0.001** |
| zHC-zFL | ASD(Ref) - TDS | 0.278±1.03 | **0.009** | 1.489 | 1.009-2.197 | 0.135 |
| | ASD(Ref) - TDP | -0.109±1.10 | 0.882 | 0.813 | 0.575-1.149 | 0.720 |
| | TDS(Ref) - TDP | -0.411±1.07 | **<0.001** | 0.502 | 0.341-0.738 | **<0.001** |
| zBPD-zFL | ASD(Ref) - TDS | 0.520±1.50 | **<0.001** | 1.482 | 1.136-1.934 | **0.012** |
| | ASD(Ref) - TDP | -0.295±1.50 | 0.111 | 0.733 | 0.562-0.955 | 0.063 |
| | TDS(Ref) - TDP | -0.858±1.50 | **<0.001** | 0.489 | 0.368-0.650 | **<0.001** |
| **Third Trimester: 116 ASD-TDS pairs, 116 ASD-TDP pairs** |  |  |  |  |  |  |
| zBPD | ASD(Ref) - TDS | 0.061±2.05 | 1 | 0.920 | 0.743-1.140 | 1 |
| | ASD(Ref) - TDP | -0.174±2.13 | 1 | 0.925 | 0.773-1.106 | 1 |
| | TDS(Ref) - TDP | -0.351±2.21 | 0.270 | 0.985 | 0.819-1.184 | 1 |
| zHC | ASD(Ref) - TDS | 0.040±1.34 | 1 | 0.924 | 0.670-1.274 | 1 |
| | ASD(Ref) - TDP | -0.028±1.34 | 1 | 0.968 | 0.729-1.287 | 1 |
| | TDS(Ref) - TDP | -0.163±1.51 | 0.738 | 0.977 | 0.749-1.275 | 1 |
| zAC | ASD(Ref) - TDS | -0.084±1.38 | 1 | 0.868 | 0.637-1.182 | 1 |
| | ASD(Ref) - TDP | -0.048±1.73 | 1 | 0.966 | 0.777-1.202 | 1 |
| | TDS(Ref) - TDP | 0.019±1.73 | 1 | 1.050 | 0.835-1.321 | 1 |
| zFL | ASD(Ref) - TDS | -0.176±1.06 | 0.237 | 0.745 | 0.502-1.106 | 0.432 |
| | ASD(Ref) - TDP | 0.082±1.09 | 1 | 1.150 | 0.813-1.627 | 1 |
| | TDS(Ref) - TDP | 0.218±1.16 | 0.135 | 1.407 | 0.994-1.992 | 0.162 |
| zBPD-zHC | ASD(Ref) - TDS | 0.039±1.37 | 1 | 0.908 | 0.651-1.266 | 1 |
| | ASD(Ref) - TDP | -0.103±1.48 | 1 | 0.910 | 0.702-1.178 | 1 |
| | TDS(Ref) - TDP | -0.167±1.54 | 0.735 | 1.022 | 0.781-1.337 | 1 |
| zHC-zAC | ASD(Ref) - TDS | 0.181±1.11 | 0.273 | 1.244 | 0.845-1.830 | 0.804 |
| | ASD(Ref) - TDP | 0.044±1.47 | 1 | 1.043 | 0.805-1.351 | 1 |
| | TDS(Ref) - TDP | -0.164±1.51 | 0.726 | 0.923 | 0.708-1.204 | 1 |
| zBPD-zAC | ASD(Ref) - TDS | 0.094±1.70 | 1 | 0.922 | 0.711-1.196 | 1 |
| | ASD(Ref) - TDP | -0.097±1.89 | 1 | 0.947 | 0.774-1.158 | 1 |
| | TDS(Ref) - TDP | -0.307±1.95 | 0.276 | 0.984 | 0.797-1.215 | 1 |
| zHC-zFL | ASD(Ref) - TDS | 0.241±1.07 | 0.063 | 1.235 | 0.829-1.841 | 0.897 |
| | ASD(Ref) - TDP | -0.148±1.10 | 0.498 | 0.782 | 0.552-1.109 | 0.504 |
| | TDS(Ref) - TDP | -0.401±1.44 | **0.009** | 0.765 | 0.574-1.018 | 0.198 |
| zBPD-zFL | ASD(Ref) - TDS | 0.201±1.88 | 0.792 | 0.986 | 0.781-1.245 | 1 |
| | ASD(Ref) - TDP | -0.284±1.85 | 0.333 | 0.845 | 0.684-1.043 | 0.348 |
| | TDS(Ref) - TDP | -0.544±2.09 | **0.018** | 0.894 | 0.734-1.088 | 0.786 |

<sup>a</sup> Paired T-test: ASD-TDS, ASD-TDP; Two-sided t-test: TDS-TDP; all p-values are Bonferroni corrected for multiple comparison (N=3)
<sup>b</sup> ASD-TDS: adjusted to maternal age and gender; ASD-TDP: adjusted to maternal age; TDS-TDP: adjusted to maternal age, gender and ethnicity.

To verify that our results were not biased by the different samples in the two trimesters (**Figure 1**), we repeated these analyses in a subsample containing only subjects with US biometric data from both trimesters. The repeat analysis confirmed the above findings, with some differences in head size between ASD and TDP becoming even clearer in this subsample (**Table 4**).

**Table 4.** Group Differences in Fetal Biometric Measures for Children with Longitudinal US Data.

| Variable | Group comparison | Mean difference ± SD | P-value<sup>a</sup> | Adjusted odds ratio (aOR)<sup>b</sup> | 95% CI | P-value<sup>a</sup> |
| --- | --- | --- | --- | --- | --- | --- |
| **Second Trimester** |  |  |  |  |  |  |
| zBPD | ASD(Ref) - TDS | 0.506±1.64 | **0.024** | 1.250 | 0.912-1.712 | 0.411 |
| | ASD(Ref) - TDP | -0.551±1.8 | **0.030** | 0.636 | 0.462-0.874 | **0.015** |
| | TDS(Ref) - TDP | -1.057±1.64 | **<0.001** | 0.463 | 0.310-0.691 | **<0.001** |
| zHC | ASD(Ref) - TDS | 0.279±1.25 | 0.153 | 1.141 | 0.756-1.722 | 1 |
| | ASD(Ref) - TDP | -0.371±1.24 | **0.039** | 0.530 | 0.338-0.832 | **0.018** |
| | TDS(Ref) - TDP | 0.650±1.21 | **<0.001** | 0.382 | 0.216-0.674 | **0.003** |
| zAC | ASD(Ref) - TDS | 0.177±1.11 | 0.492 | 1.060 | 0.669-1.680 | 1 |
| | ASD(Ref) - TDP | -0.319±1.37 | 0.159 | 0.664 | 0.440-1.003 | 0.156 |
| | TDS(Ref) - TDP | -0.496±1.31 | **0.009** | 0.569 | 0.330-0.981 | 0.129 |
| zFL | ASD(Ref) - TDS | 0.082±1.06 | 1 | 1.033 | 0.653-1.636 | 1 |
| | ASD(Ref) - TDP | 0.053±1.07 | 1 | 1.140 | 0.710-1.829 | 1 |
| | TDS(Ref) - TDP | -0.029±1.00 | 1 | 1.019 | 0.594-1.749 | 1 |
| zBPD-zHC | ASD(Ref) - TDS | 0.255±1.09 | 0.126 | 1.425 | 0.907-2.239 | 0.375 |
| | ASD(Ref) - TDP | -0.199±1.08 | 0.381 | 0.686 | 0.421-1.116 | 0.387 |
| | TDS(Ref) - TDP | -0.454±1.16 | **0.009** | 0.552 | 0.340-0.895 | **0.048** |
| zHC-zAC | ASD(Ref) - TDS | 0.104±1.04 | 1 | 1.120 | 0.703-1.782 | 1 |
| | ASD(Ref) - TDP | -0.039±1.28 | 1 | 0.911 | 0.610-1.359 | 1 |
| | TDS(Ref) - TDP | -0.143±1.16 | 1 | 0.756 | 0.470-1.214 | 0.741 |
| zBPD-zAC | ASD(Ref) - TDS | 0.354±1.43 | 0.093 | 1.300 | 0.920-1.836 | 0.411 |
| | ASD(Ref) - TDP | -0.244±1.63 | 0.630 | 0.797 | 0.577-1.100 | 0.504 |
| | TDS(Ref) - TDP | -0.598±1.53 | **0.009** | 0.610 | 0.423-0.881 | **0.024** |
| zHC-zFL | ASD(Ref) - TDS | 0.194±1.02 | 0.288 | 1.164 | 0.706-1.919 | 1 |
| | ASD(Ref) - TDP | -0.417±1.07 | **0.003** | 0.378 | 0.219-0.654 | **0.003** |
| | TDS(Ref) - TDP | -0.611±1.01 | **<0.001** | 0.283 | 0.145-0.552 | **<0.001** |
| zBPD-zFL | ASD(Ref) - TDS | 0.449±1.42 | 0.018 | 1.344 | 0.942-1.917 | 0.309 |
| | ASD(Ref) - TDP | -0.616±1.57 | **0.003** | 0.520 | 0.357-0.757 | **0.003** |
| | TDS(Ref) - TDP | -1.065±1.48 | **<0.001** | 0.380 | 0.238-0.608 | **<0.001** |
| **Third Trimester** |  |  |  |  |  |  |
| zBPD | ASD(Ref) - TDS | 0.029±2.30 | 1 | 0.851 | 0.671-1.080 | 0.552 |
| | ASD(Ref) - TDP | -0.579±2.45 | 0.159 | 0.789 | 0.631-0.988 | 0.117 |
| | TDS(Ref) - TDP | -0.608±2.32 | 0.120 | 0.876 | 0.681-1.127 | 0.912 |
| zHC | ASD(Ref) - TDS | 0.144±1.42 | 1 | 0.893 | 0.613-1.301 | 1 |
| | ASD(Ref) - TDP | -0.269±1.34 | 0.294 | 0.725 | 0.483-1.088 | 0.363 |
| | TDS(Ref) - TDP | -0.414±1.49 | 0.093 | 0.755 | 0.514-1.108 | 0.450 |
| zAC | ASD(Ref) - TDS | 0.056±1.46 | 1 | 0.915 | 0.647-1.294 | 1 |
| | ASD(Ref) - TDP | -0.008±1.53 | 1 | 0.965 | 0.686-1.356 | 1 |
| | TDS(Ref) - TDP | -0.064±1.68 | 1 | 0.990 | 0.709-1.383 | 1 |
| zFL | ASD(Ref) - TDS | -0.056±1.04 | 1 | 0.781 | 0.485-1.259 | 0.933 |
| | ASD(Ref) - TDP | -0.007±1.05 | 1 | 0.958 | 0.591-1.551 | 1 |
| | TDS(Ref) - TDP | 0.049±1.13 | 1 | 1.121 | 0.695-1.808 | 1 |
| zBPD-zHC | ASD(Ref) - TDS | -0.066±1.61 | 1 | 0.839 | 0.605-1.162 | 0.873 |
| | ASD(Ref) - TDP | -0.307±1.66 | 0.402 | 0.746 | 0.537-1.038 | 0.249 |
| | TDS(Ref) - TDP | -0.240±1.64 | 0.753 | 0.968 | 0.680-1.376 | 1 |
| zHC-zAC | ASD(Ref) - TDS | 0.081±1.26 | 1 | 0.994 | 0.669-1.477 | 1 |
| | ASD(Ref) - TDP | -0.266±1.25 | 0.246 | 0.732 | 0.479-1.118 | 0.447 |
| | TDS(Ref) - TDP | -0.347±1.45 | 0.183 | 0.735 | 0.497-1.086 | 0.366 |
| zBPD-zAC | ASD(Ref) - TDS | -0.069±1.88 | 1 | 0.824 | 0.624-1.089 | 0.522 |
| | ASD(Ref) - TDP | -0.579±1.95 | **0.048** | 0.709 | 0.534-0.943 | 0.054 |
| | TDS(Ref) - TDP | -0.510±1.79 | 0.078 | 0.832 | 0.600-1.155 | 0.816 |
| zHC-zFL | ASD(Ref) - TDS | 0.236±1.33 | 0.378 | 1.102 | 0.748-1.624 | 1 |
| | ASD(Ref) - TDP | -0.272±1.20 | 0.192 | 0.691 | 0.440-1.087 | 0.330 |
| | TDS(Ref) - TDP | -0.509±1.40 | **0.015** | 0.640 | 0.421-0.972 | 0.108 |
| zBPD-zFL | ASD(Ref) - TDS | 0.081±2.18 | 1 | 0.898 | 0.707-1.142 | 1 |
| | ASD(Ref) - TDP | -0.585±2.20 | 0.090 | 0.751 | 0.584-0.966 | 0.078 |
| | TDS(Ref) - TDP | -0.666±2.10 | **0.039** | 0.820 | 0.623-1.079 | 0.471 |

<sup>a</sup> Paired T-test: ASD-TDS, ASD-TDP; Two-sided t-test: TDS-TDP; all p-values are Bonferroni corrected for multiple comparison (N=3).
<sup>b</sup> ASD-TDS: adjusted to maternal age and gender; ASD-TDP: adjusted to maternal age; TDS-TDP: adjusted to maternal age, gender and ethnicity.

Head biometric measures were consistently smaller for females than males within and across groups (**Figures 2-4; Table 5**), indicating a significant sex effect in fetal growth measures. Therefore, we reevaluated the group differences in biometric measures separately for males and females (**Table 6**). In the second trimester, the BPD in both ASD and TDS males was significantly smaller compared to TDP, but no such differences were seen in HC measures. As a result, ASD and TDS males had narrower and more elongated heads compared to TDP males (zBPD-zHC=-0.271±1.04; p=0.027 and zBPD-zHC=-0.539±1.08; p<0.001 for ASD-TDP and TDS-TDP, respectively). In the third trimester, the differences in head size and shape were attenuated and became statistically insignificant. In females, HC and BPD were proportionally smaller in ASD and TDS compared to TDP in both pregnancy trimesters (**Table 6**), thus suggesting that such head shape abnormality (narrower and elongated) during mid gestation is an exclusive characteristic of males in ASD families.

**Figure 4:**
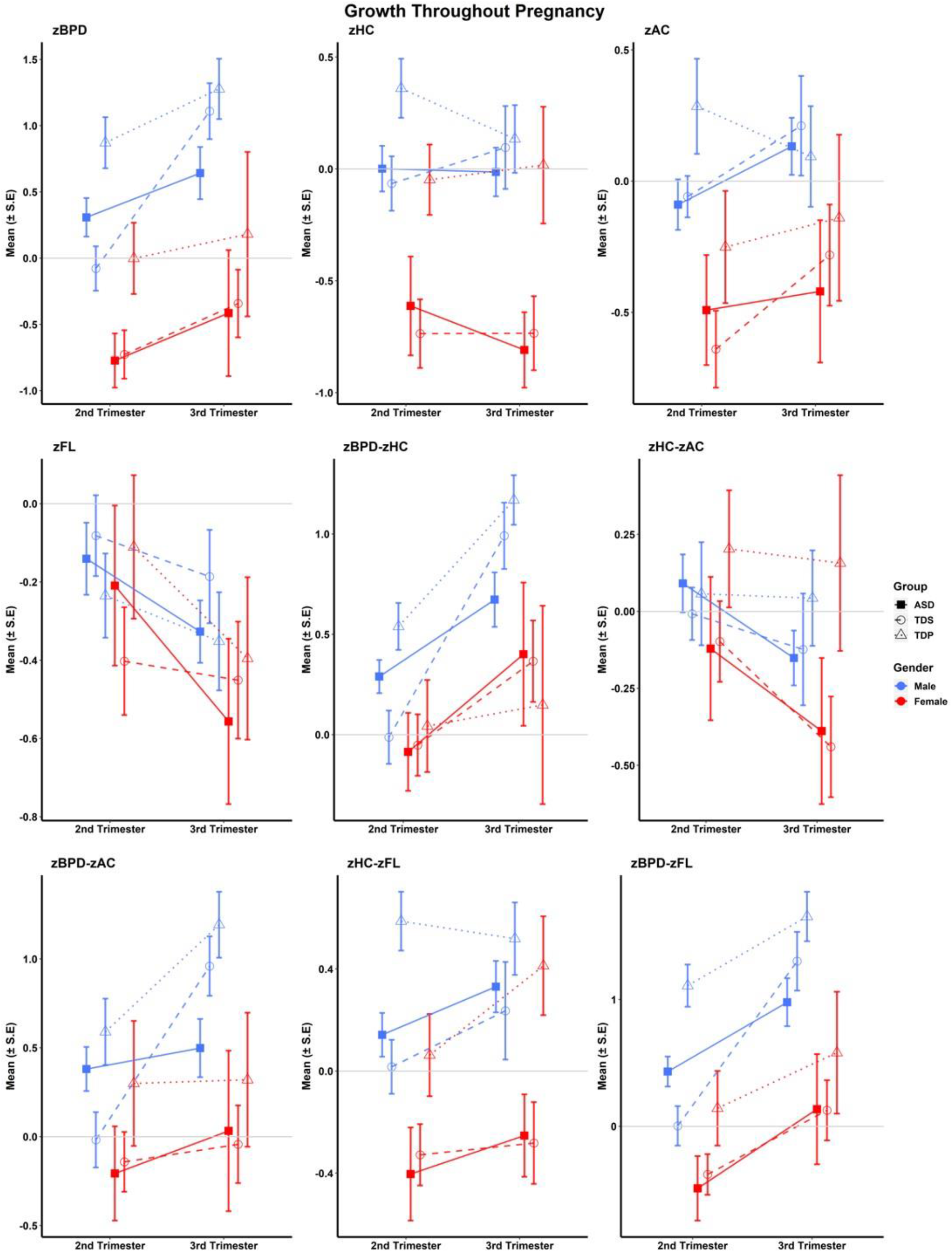
Changes in biometric measures between second and third trimesters. The mean and SE of different biometric measures are shown for males (blue) and females (red) in the three study groups: ASD (squares), TDS (empty circles) and TDP (empty diamonds). A steeper line between the two trimesters indicates a faster growth rate between the two trimesters.

**Table 5.** Sex Differences in Biometric Measures.

| Variable | Male mean (SD) | Female mean (SD) | P-value<sup>a</sup> |
| --- | --- | --- | --- |
| **Second Trimester: 277 males, 106 females** |  |  |  |
| zBPD | 0.408(1.26) | -0.475(1.15) | **<0.001** |
| zHC | 0.093(0.88) | -0.479(0.88) | **<0.001** |
| zAC | -0.018(.87) | -0.507(0.83) | **<0.001** |
| zFL | -0.127(0.75) | -0.262(0.84) | 0.127 |
| zBPD-zHC | 0.309(0.78) | 0.026(0.77) | **0.002** |
| zHC-zAC | 0.116(0.79) | 0.036(0.77) | 0.403 |
| zBPD-zAC | 0.424(1.07) | 0.045(1.04) | **0.002** |
| zHC-zFL | 0.220(0.74) | -0.226(0.81) | **<0.001** |
| zBPD-zFL | 0.529(1.10) | -0.210(1.09) | **<0.001** |
| **Third Trimester: 256 males, 99 females** |  |  |  |
| zBPD | 0.851(1.48) | -0.118(1.71) | **<0.001** |
| zHC | 0.030(1.03) | -0.488(0.96) | **<0.001** |
| zAC | 0.052(1.19) | -0.152(1.05) | 0.135 |
| zFL | -0.324(0.79) | -0.341(0.86) | 0.861 |
| zBPD-zHC | 0.831(0.99) | 0.366(1.25) | **0.001** |
| zHC-zAC | -0.017(1.01) | -0.329(0.91) | **0.008** |
| zBPD-zAC | 0.835(1.34) | 0.052(1.37) | **<0.001** |
| zHC-zFL | 0.361(0.99) | -0.144(0.77) | **<0.001** |
| zBPD-zFL | 1.196(1.45) | 0.235(1.42) | **<0.001** |

<sup>a</sup> Two-sided t-test

**Table 6.**
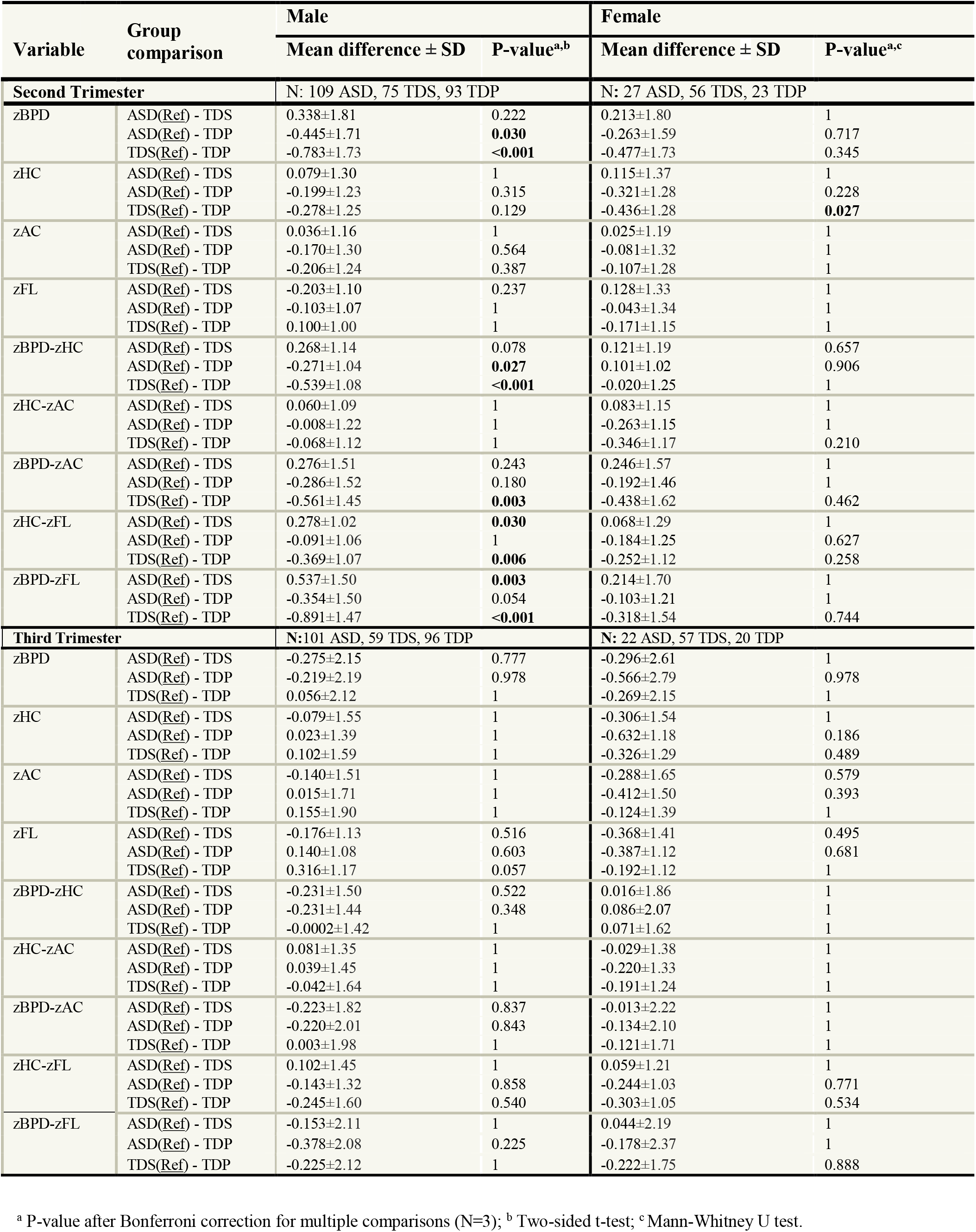
Group Differences in Fetal Biometric Measures for Males and Females.

### Longitudinal Growth

For approximately half the ASD fetuses and one-third of the controls there were biometric measures for both the second and third trimesters (**Figure 1**). These data allowed us to examine differential rates of organ growth during pregnancy (**Figure 4; Table 7**). Head width (zBPD) showed the most considerable growth compared to other body parts in both cases and controls and in both males and females. However, TDS males exhibited significantly faster BPD growth than both ASD and TDP males (β=0.084 and β=0.100 respectively; p<0.001). As a result, the gap between the TDS and TDP groups in the second trimester closed in the third trimester, making the head width of male fetuses later diagnosed with ASD smaller in the third trimester vs both control groups.

**Table 7.** Growth Rate Throughout Pregnancy in Male and Female Fetuses.

| Variable | Group comparison<sup>a</sup> | Male |  |  | Female |  |  |
| --- | --- | --- | --- | --- | --- | --- | --- |
| | | Adjusted $\beta^b$ | 95% CI | P-value<sup>c</sup> | Adjusted $\beta^b$ | 95% CI | P-value<sup>c</sup> |
| zBPD | ASD(Ref) - TDS | 0.084 | 0.041-0.127 | **<0.001** | -0.010 | -0.087-0.067 | 1 |
|  | ASD(Ref) - TDP | <0.001 | -0.038-0.038 | 1 | -0.056 | -0.166-0.055 | 0.951 |
|  | TDP(Ref) - TDS | 0.100 | 0.052-0.147 | **<0.001** | 0.030 | -0.057-0.116 | 1 |
| zHC | ASD(Ref) - TDS | 0.013 | -0.017-0.043 | 1 | 0.022 | -0.032-0.077 | 1 |
|  | ASD(Ref) - TDP | -0.035 | -0.063- -0.007 | **0.045** | 0.035 | -0.026-0.096 | 0.783 |
|  | TDP(Ref) - TDS | 0.092 | 0.041-0.143 | **0.012** | -0.009 | -0.067-0.049 | 1 |
| zAC | ASD(Ref) - TDS | <-0.001 | -0.028-0.028 | 1 | 0.017 | -0.021-0.056 | 1 |
|  | ASD(Ref) - TDP | -0.043 | -0.071- -0.014 | **0.009** | -0.013 | -0.078-0.053 | 1 |
|  | TDP(Ref) - TDS | 0.054 | 0.013-0.095 | **0.030** | 0.033 | -0.017-0.084 | 0.549 |
| zFL | ASD(Ref) - TDS | -0.001 | -0.024-0.023 | 1 | -0.008 | -0.046-0.030 | 1 |
|  | ASD(Ref) - TDP | -0.003 | -0.027-0.021 | 1 | -0.005 | -0.059-0.050 | 1 |
|  | TDP(Ref) - TDS | -0.007 | -0.033-0.020 | 0.617 | -0.001 | -0.047-0.044 | 1 |
| zBPD-zHC | ASD(Ref) - TDS | 0.025 | 0.006-0.044 | **0.018** | -0.004 | -0.060-0.052 | 1 |
|  | ASD(Ref) - TDP | 0.024 | -0.005-0.053 | 0.318 | -0.077 | -0.165-0.011 | 0.252 |
|  | TDP(Ref) - TDS | 0.031 | -0.093-0.155 | 0.687 | 0.084 | 0.012-0.156 | 0.069 |
| zHC-zAC | ASD(Ref) - TDS | -0.002 | -0.036-0.032 | 1 | -0.052 | -0.087—0.017 | **0.012** |
|  | ASD(Ref) - TDP | 0.023 | -0.012-0.058 | 0.576 | 0.057 | -0.013-0.128 | 0.318 |
|  | TDP(Ref) - TDS | -0.038 | -0.085-0.008 | 0.312 | -0.032 | -0.086-0.021 | 0.699 |
| zBPD-zAC | ASD(Ref) - TDS | 0.089 | 0.046-0.132 | **<0.001** | 0.021 | -0.050-0.092 | 1 |
|  | ASD(Ref) - TDP | 0.001 | -0.037-0.039 | 1 | -0.044 | -0.147-0.060 | 1 |
|  | TDP(Ref) - TDS | 0.041 | -0.011-0.094 | 0.366 | 0.056 | -0.018-0.130 | 0.384 |
| zHC-zFL | ASD(Ref) - TDS | 0.014 | -0.018-0.046 | 1 | 0.029 | -0.002-0.060 | 0.201 |
|  | ASD(Ref) - TDP | -0.025 | -0.055-0.006 | 0.324 | 0.051 | -0.0004-0.103 | 0.156 |
|  | TDP(Ref) - TDS | 0.039 | 0.005-0.074 | 0.081 | -0.017 | -0.072-0.038 | 1 |
| zBPD-zFL | ASD(Ref) - TDS | 0.095 | 0.051-0.138 | **<0.001** | 0.002 | -0.075-0.078 | 1 |
|  | ASD(Ref) - TDP | 0.001 | -0.037-0.039 | 1 | -0.044 | -0.147-0.060 | 1 |
|  | TDP(Ref) - TDS | 0.088 | 0.041-0.135 | **<0.001** | 0.031 | -0.049-0.112 | 1 |

<sup>a</sup> **85 ASD** (70 males, 15 females), **69 TDS** (36 males, 33 females), **56 TDP** (43 males, 13 females).
<sup>b</sup> Interaction of ASD diagnosis with pregnancy week assessed by linear mixed effect regression adjusted to ethnicity. Positive $\beta$ means faster growth rate compared to reference group.
<sup>c</sup> P-value after Bonferroni correction for multiple comparisons (N=3).

### Association with ASD Phenotypes

Finally, we examined the association between fetal biometric measures and the severity of ASD symptoms (**Figure 5**). Three major trends were seen in this analyses: 1) smaller biometric measures (i.e. HC, BPD, AC, and FL) were associated with worse ASD symptoms especially among males in the second trimester (Figure **5A**) and females in third trimester (**Figure 5D**); 2) larger head-to-body ratios were positively correlated with more severe ASD symptoms, especially in the second trimester in both males and females (**Figures 5A and 5B** respectively); and 3) In females, different trends were seen between the two trimesters. In the second trimester, most biometric measures were positively correlated with ASD severity (**Figure 5C**), while in the third trimester, they were inversely correlated with symptom severity (**Figure 5D**). Of note, none of these correlations remained statistically significant after Bonferroni correction for multiple testing.

**Figure 5:**
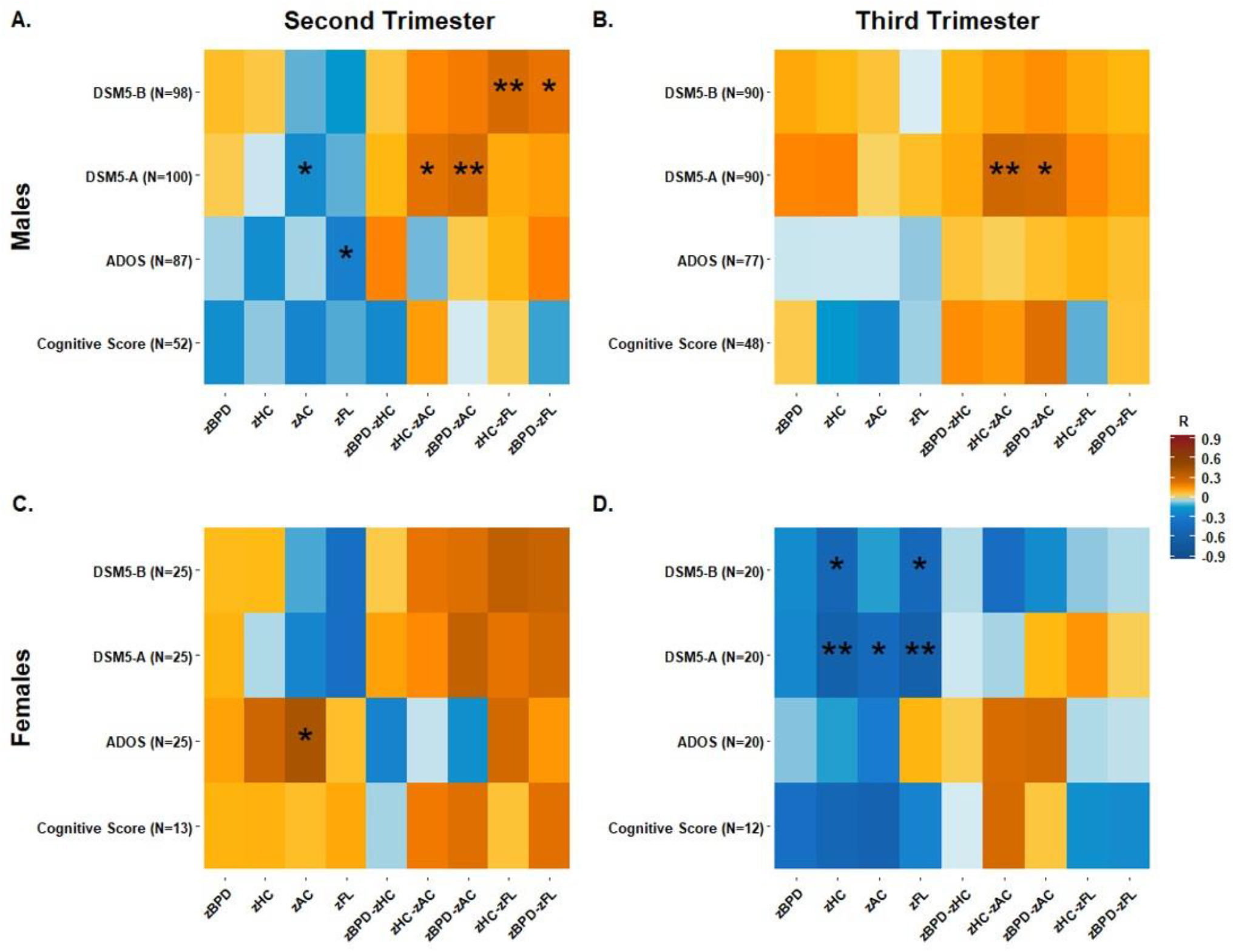
Correlation between ASD Severity and Fetal Biometric Measures in the Second and Third Trimesters of pregnancy in Males and Females. Spearman correlation coefficients (R) are color coded, with asterisks indicating statistically significant correlations: * P-value<0.05, ** P-value≤0.01. For all tests, a positive R means a larger biometric measure is correlated with worse outcome of specific clinical test, as follows:DSM5-B: Restrictive-Repetitive Domain has a range of three values (1) Requiring Support, (2) Requiring Substantial Support, (3) Requiring Very Substantial Support. A higher score means a worse outcome. DSM5-A: Social Domain has a range of three values (1) Requiring Support, (2) Requiring Substantial Support, (3) Requiring Very Substantial Support. A higher score means worse outcome. ADOS: A higher score means a worse outcome. Cognitive Score: Cognitive abilities of child. In the original test. a higher score means better outcome. In order to avoid confusion, we multiplied correlation outcome by -1 so that in this test too, a positive correlation means a worse outcome.

## Discussion

This is the largest and most comprehensive study to date to examine the association between fetal growth parameters and ASD. Three main findings (discussed in detail below) emerged from this study. First, fetuses later diagnosed with ASD and their TDS have narrower heads during mid-gestation compared to TDP, thus suggesting that such fetal head growth abnormality is a familial trait of ASD. Second, ASD-related head growth abnormalities are modulated by the sex of the fetus, with males and females showing different head shapes during gestation. Third, fetal head anomalies appear to be associated with the severity of ASD.

The abnormal fetal head growth associated with ASD that we report in this study suggests that brain abnormalities associated with ASD begin during gestation. Further support for prenatal brain anomalies in ASD can be found in a recent review which summarizes findings from multiple studies of ASD about abnormal prenatal development of the brain including cell proliferation, neurogenesis, migration, laminar organization, and neurite outgrowth during the first and second trimester alongside neurite outgrowth, synaptogenesis, and synapse functioning in the third trimester and early postnatal life.^10^ Additional support may be drawn from postmortem studies that found clues of aberrant neuronal migration during early prenatal brain development in the prefrontal and temporal cortical tissue, alongside evidence of curtailment of maturation of the forebrain limbic system and abnormalities in the cerebellar circuits, all of which are brain regions held to be affected ASD.^5,6,8,9^ In addition, two postnatal MRI studies in preterm infants indicated structural brain asymmetry and reduced brain volume in preterm fetuses later diagnosed with ASD, again suggesting that abnormal head growth begins early in prenatal life, though ASD in the context of extreme prematurity might have a different pathogenic pathway compared to ASD in the general population.^54,55^ Moreover, a recent MRI study found a 15% increase of extra-axial CSF volume among ASD children during early childhood.^19^ This increase is categorised as benign external hydrocephalus, characterized by increased CSF mainley above the frontal lobes, and is suggested to accure due to delayed maturation of arachnoid granulations which leads to an imbalance between the production and absorption of CSF.^56^ Our finding of a narrow and elongated head among ASD males is consistent with this fingdings, and may suggest the increase of CSF above the frontal lobes may begin during prenatal life due to the delayed maturation of arachnoid granulations, leading to the abnormal and elongated head shape.

Our findings suggest that abnormalities in the fetal head during gestation that are later associated with ASD is a familial trait. Support for this premise can be drawn both from a prospective study showing smaller fetal heads in families with ASD compared to fetuses from families without ASD^36^ and from a study that found an association between smaller HC and various autistic traits^37^, which are more prevalent in ASD families.^57^ Additional support comes from genetic studies suggesting that HC is highly heritable^58^ and that genetic variations associated with HC are also associated with risk of ASD.^59^ Another indication of a familial link between abnormal fetal head growth and risk of ASD lies in some maternal endocrine functions during pregnancy that have been associated with both of these traits.^60,61^ Interestingly, fetuses of our TDS group demonstrated accelerated head growth during late gestation to compensate for their relatively smaller heads, a finding consistent with previous study demonstrating slightly faster growth rate among TDS compared to TDP.^36^ Similar accelerated head growth in early infancy has been reported in ASD children,^7,11-15^ providing another familial trait of head growth abnormality associated with ASD. Furthermore, adjustment of brain developmental rate has also been shown as type of whole-brain adaptation in response to early environmental adversity in order to maximize its fit with external enviorment.^62^

Our findings also suggest that ASD is associated with different prenatal head growth abnormalities in males and females. Similar sex-specific head size abnormalities were seen in a large population-based study at birth and throughout early childhood in children with ASD.^18^ In addition, differences in total brain volume, and in gray and white matter volumes in different brain regions were seen throughout childhood in boys and girls with ASD.^14,16,18,22,28-30^ Furthermore, our study suggests that the observed sex-specific prenatal head growth abnormalities in fetuses later diagnosed with ASD are correlated with symptom severity in both sexes. This finding is consistent with previous postnatal studies that reported correlations between postnatal head size, brain morphometry, and extra-axial CSF volume on the one hand, to behavioral development, IQ,^21^ social deficits,^15^ sleep disturbances, lower non-verbal ability, poorer motor ability,^19^ and ADOS scores,^22^ on the other. We therefore suggest that postnatal sex-specific differences in ASD severity are associated with differences in fetal brain development between males and females later diagnosed with ASD.

Our study has several advantages over previous investigations of prenatal US data of children with ASD. The use of two distinct control groups, TDS and TDP, enabled us to adjust our findings to multiple familial and prenatal confounders that are known to have a considerable effect on both ASD risk and fetal growth (e.g., sex^26,40^, shared genetics among siblings^1,2,38,39^), making our findings more compelling. In this context, previous findings suggesting larger HC^34^ or BPD^33^ in ASD fetuses may be attributed to bias resulting from the large and unaddressed proportion of male cases (~90% in both), for which biometric measures are usually larger.^40^ Another important advantage of our study is the availability of longitudinal biometric measures from both second and third trimesters for a significant portion of our sample, thus allowing us to account for sampling bias between the two pregnancy periods and to investigate prenatal growth trajectories of these biometric measures in ASD and the two control groups. Using this data, we show that significant head growth abnormalities that are seen in mid-gestation (second trimester) for the ASD and TDS groups are attenuated towards the end of the pregnancy. While this attenuation could be due to accelerated growth in some fetuses like is seen for the TDS males, it could also be attributed to the typical higher variability of gestation-based biometric measures during the third trimester.^48^ Finally, this study is the first to examine correlations between prenatal US measures and ASD clinical metrics, allowing us to understand the associations between fetal growth and ASD severity.

Our study has several limitations. First, the sample size of the study, despite being the largest of its kind to date, is still too small to address associations within subgroups. This is particularly relevant to the longitudinal analyses (**Figure 4 & table 7**) and to the correlations between the biometric measures and ASD symptoms (**Figure 5**) where data was available only for subsets of our sample. The same limitation applies to the sex-specific analyses since females comprised only ~20% of the cases due to the inherent male bias of ASD.^26^ In addition, parents of children in the study sample were slightly younger than those of the other children with ASD in the NARC cohort. However, this small difference in parental age is unlikely to affect the study findings. Finally, we had only limited data about potential birth and pregnancy confounding factors for subjects in our study, such as weight at birth. Nonetheless, the comparison of cases to both TDS and TDP control groups probably minimized the effect of such confounders.

## Conclusions

Our findings suggest that abnormal fetal head growth is a familial trait of ASD, which is modulated by sex and is associated with the severity of the disorder. Thus, it could serve as an early biomarker for ASD. Integration of our findings with fetal brain research will shed important light on neuroanatomical embryonic milestones in ASD development.

## Data Availability

Data about ASD status and characteristics were obtained from the database of the National Autism Research Center of Israel

https://www.autismisrael.org/regional-database

## Acknowledgements

This study was supported by a grant from the Israeli Science Foundation (527/15).

This study was conducted as part of the requirements to obtain a degree in medicine from the Joyce & Irving

Goldman Medical School, Faculty of Health Sciences, Ben-Gurion University of the Negev.

We thank Mrs. Inez Mureink and Prof. Norm O’Rourke for critical reviewing and editing of the manuscript.

